# Plant Formulation ATRICOV 452 in Improving the Level of COVID-19 Specific Inflammatory Markers in Patients

**DOI:** 10.1101/2021.10.07.21264491

**Authors:** Latha Damle, Hrishikesh Damle, BR Bharath

## Abstract

Due to the huge demand for health care facilities, there is a need for safe therapeutic intervention which can reduce the need for extensive health care support. In that regard, the current study was aimed at performing Phase 1 clinical trial to determine the safety of plant formulation in 24 healthy volunteers and Phase 2 trials in 100 COVID-19 patients to determine the tolerability and impact on the level of COVID 19 specific inflammatory markers. The outcomes of the Phase 1study have suggested the safe usage of plant formulation in humans and encouraged to conduct Phase 2 clinical trial. In the Phase 2 trial, the plant formulation was evaluated in 100 COVID-19 patients along with the standard of care. In Phase 1 single dose of 500 mg plant formulation capsule was used as an intervention, while 1gm thrice a day of plant formulation for 14 days was the testing dose for Phase 2. During the Phase 1 trial, no adverse event was observed and all organ systems were normal in function. During the Phase 2 trial, 100 patients underwent randomization, 50 were assigned to receive plant formulation, and 50 to receive placebo. Three patients in the placebo and two patients in the plant formulation group had dropped out from the study. Hence, the primary analysis population included 95 patients (48 allocated to plant formulation and 47 to placebo). The COVID 19 specific inflammatory markers improved faster and became normal in the plant formulation treatment group. In conclusion, the plant formulation (ATRICOV 452) has been found to be safe in phases 1 and 2.

**Trial Registration:** Phase 1: CTRI Registration number: CTRI/2020/09/027660

Phase 2: CTRI Registration number: CTRI/2021/01/030795

## 1. Introduction

During this growing COVID-19 pandemic situation, as of September 30th, 2020, the World Health Organization (WHO) has reported that 33,502,430 and 1,004,421 death cases have been confirmed worldwide, and it has spread to 235 countries and the demand for supportive care facilities is also increasing (WHO, 2020). Currently, there is no effective cure for SARS-CoV-2 infection and the most common treatment for patients with COVID-19 is supportive care (Sanders et al., 2020). Meanwhile, the existing supportive care facility for COVID-19 treatment is insufficient to anticipate the growing demand (Jason et al., 2020). Hence, and efforts are being made globally towards identifying the need for supportive care on the basis of severity, spectrum, and impact of the disease and extend the facility to the needy. Multiple repurposed anti-viral drugs, including remdesivir and lopinavir plus ritonavir, have been used as adjunct drugs in COVID-19 treatment and their intravenous administration requires the clinician’s assistance (Wang et al. 2020; Li and De Clercq 2020; Chen et al., 2020). As SARS-CoV-2 is highly contagious, that can be transmitted from person to person in a hospital with high traffic. Hence, reducing hospital visits and hospitalisation by using oral anti-SARS-CoV-2 drugs is also an option to reduce traffic in hospitals (Melika et al., 2020;). In that regard, effective strategies for prophylaxis and holistic management are of paramount importance in curtailing the progress of the disease and reducing the burden on hospitals.

In this study, phase 1 and phase 2 clinical trials were conducted to evaluate the safety and tolerability of the plant formulation ATRICOV 452 which is consisting of equal proportion of plant extracts (Latha et al., 2021). The ATRICOV 452 was oral administered to 24 healthy volunteers and 100 COVID-19 patients in phase 1 and phase 2 respectively.

## 2. Materials and Methods

### 2.1. Phase 1 clinical trial

In spite of known safety, the CTRI approved Phase-1 study entitled “A Prospective, Interventional, Open-label, Phase 1, single-center, single fasting dose study to evaluate the safety and tolerability of ATRICOV 452 capsule of 500 mg in healthy adult human subjects” was conducted in the CDSO-DCGI approved CRO ICBio Clinical Research Pvt. Ltd., Bangalore, located at Bangalore. The study was interventional, single fasting dose, single-arm with the objective of assigning the safety of plant formulation (ATRICOV 452) in 24 healthy volunteers. The study protocol was approved by an in-house ethical committee constituted by CRO and conducted from 17 Sep 2020 to 24 Sep. 2020.

### 2.2. Phase 2 clinical trial

The single-blind, placebo-controlled, randomized clinical trial was designed to evaluate the tolerability of plant formulation (ATRICOV 452) its impact on the level of COVID-19 specific inflammatory markers in adult COVID-19 patients under standard of care also. The study was conducted in compliance with the Declaration of Helsinki,13 the Good Clinical Practice ICH guidelines, and local regulatory requirements.

#### 2.2.a. Study Design

This trial was conducted for 14 Days in two different trial sites namely, Vedant Hospital, Thane and Life point Multispeciality Hospital, Pune. The IEC approval for the study was obtained from Vedant Hospital Institutional Ethics Committee (VHIEC) and Lifepoint Research Ethics committee on January 13, 2021 and March 13, 2021. The enrolment process was started from March 19, 2021, to June 16, 2021 without regard to sex, race, ethnicity, or religion. Patients were randomized based on stratified randomization method where, the randomization was based on mild or moderate symptoms and presence of co-morbidity like diabetes & hypertension over active and placebo arm. Diagnosis of SARS-CoV-2 infection was done using RT-PCR and COVID-19 (Blood test: Total leucocyte count, NLR. CRP, Ferritin, D-Dimer, LOH, IL-6, PCT and ESR, Chest X-Ray and CT-Scan) specific investigations were carried out to assess the severity of the disease. Medical history, Clinical examination, Vital signs were recorded carefully. The patients were classified into mild, Moderate and severe according to following criteria: Patients with uncomplicated upper respiratory tract infection possessing SpO2 ≥94% in room air, RR ≤24/m and no evidence of hypoxemia or breathlessness were considered as mild. The patients with pneumonia with no signs of severe disease, SpO2 within the range of 94%-90% in room air RR in the range of 24-30/m were considered as moderate and the patients with severe pneumonia with SpO2 < 90% in room air, RR >30/m were considered severe and not recruited for the study. The subjects (100 patients) recruited for the study were grouped into Group A and B using randomization table. Group A was given standard of care along with placebo and Group B was given standard of care along with plant formulation (ATRICOV 452). Bitterness in mouth, abdominal pain and nausea were identified as expected adverse drug reactions associated with plant formulation (ATRICOV 452). Patients with co-morbidity were treated with concomitant treatment. Periodic investigations were made as listed in schedule of events (**Table 1**). The patients were discharged as per the protocol and monitored for sign and symptoms over telephone.

**Table 1:** Schedule of events detailing the investigations performed during different treatment visits

|  | Screening | Treatment visits |  |  |  |  |  |  |  | FV<sup>^</sup> | UV<sup>^</sup> |
| --- | --- | --- | --- | --- | --- | --- | --- | --- | --- | --- | --- |
| Visit | 1 | 2 | 3 | 4 | 5 | 6 | 7 | 8 | 9 | 10 |  |
| Screening Day | 0 | 1 | 4 | 5 | 6 | 7 | 8 | 11 | 15 | 23±2 |  |
| Obtaining Informed Consent | X |  |  |  |  |  |  |  |  |  |  |
| Demography | X |  |  |  |  |  |  |  |  |  |  |
| Medical/Surgical History | X |  |  |  |  |  |  |  |  |  |  |
| Chest X-ray/ CT scan | X |  |  |  |  |  |  |  | X |  |  |
| Pregnancy test (if adult female) | X |  |  |  |  |  |  |  |  |  |  |
| Covid-19 specific Lab investigation | X |  |  |  |  | X |  | X | X |  |  |
| Covid-19 related laboratory test (IgG) | X |  |  |  |  |  |  |  |  |  |  |
| Routine Lab investigation | X |  |  |  |  |  |  |  | X |  |  |
| Vital Signs (BP, Pulse rate, Spo2/RR/Temp) | X | X | X | X | X | X | X | X | X | X | X |
| Physical Examination | X | X | X | X | X | X | X | X | X | X | X |
| Covid-19 related common sign and symptoms | X | X | X | X | X | X | X | X | X | X | X |
| Investigator assessments | X | X | X | X | X | X | X | X | X | X | X |
| Inclusion/Exclusion Criteria | X |  |  |  |  |  |  |  |  |  |  |
| IP Dispensing |  | X | X | X | X | X | X | X |  |  |  |
| Issue Dairy card |  | X | X | X | X | X | X | X |  |  |  |
| Dosing |  | X | X | X | X | X | X | X | X |  |  |
| Checking for concomitant medications (if any) |  | X | X | X | X | X | X | X | X | X | X |
| Compliance check |  | X | X | X | X | X | X | X | X |  |  |
| AE/SAE assessment monitoring |  | X | X | X | X | X | X | X | X | X | X |

<sup>^</sup> Follow Up visit , Unscheduled Visit, \* Chest X-Ray/CT if suggested by the Physician # RT PCR positive patients selected for study

#### 2.2.b. Participants

The study included adults with SARS-CoV-2 infection confirmed by RT-PCR. The patients were excluded if the physician finds vulnerability based on the history, nutritional status, physical appearance, or any other reason and did not comply with any of the inclusion and exclusion criteria (**Figure 1**).

**Figure 1:**
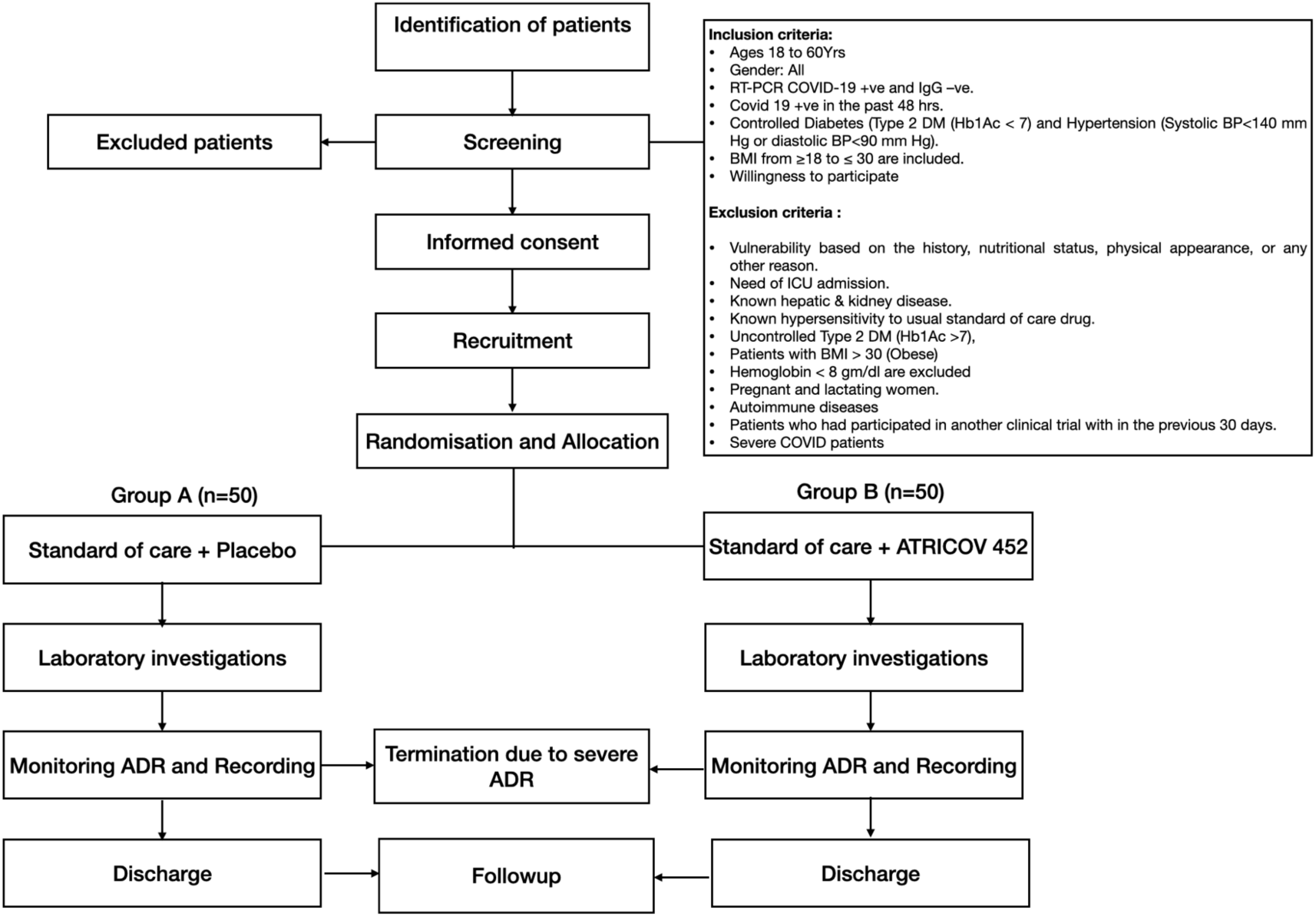
Flowchart illustrating the trial protocol, inclusion and exclusion criteria.

#### 2.2.c. Randomization and stratification

Patients were randomized 1:1 to ATRICOV 452 or identical placebo capsules. Randomization schedules were generated that stratified by signs and symptoms (mild and moderate) and co-morbidity (present and absent). Treatments were randomly allocated using randomisation table. All outcome assessors, investigators, and research staff who were involved in the trial were blinded to participant treatment assignment. The trial was carried out as per the flowchart (**Figure 1**)

#### 2.2.d. Intervention

Group A was given standard of care along with placebo containing inert starch (Identical looking 500mg) 2 capsules thrice a day for 14 days. Group B was given standard of care along with plant formulation (ATRICOV 452) (500mg) 2 capsules thrice a day for 14 days. The standard of care for mild patients was Tab-Favipiravir 1800mg 1-0-1 on Day I and 800mg 1-0-1 for 6 days (total 7 days). The standard of care for moderate patients was Oseltamavir 75mg Cap 1-0-1 for 5 days. This dose range was determined based on the preclinical and phase 1 findings.

## 3. Results and Discussion

### 3.1. Clinical trial Phase 1

The close observation was performed for safety parameters on day 5,6 and 7 after administration of ATRICOV 452 through IP mode. As per the clinical assessment, there were no allergic reactions, neurological and musculoskeletal abnormality observed during the study period. Subjects have not experienced any gastrointestinal disorders during the study period.

The results of the T-Test revealed that there was no statistically significant difference in the difference between means (p = 0.3403 and p = 0.0103) for SGOT and ALPI.

The results of the Wilcoxon signed-rank-test revealed that there was a no statistically significant difference between Means (p = 0.2373 and p = 0.3594) for SGPT and Bilirubin.

The Means of Vital Parameters (Pulse Rate, Blood Pressure, and Temperature) for different time points were not normally distributed and did not follow the assumptions for repeated measures One-way ANOVA, non-parametric mixed methods ANOVA was used to compare the Means overall different time point.

The results of all vital parameters by using non-parametric oneway repeated measures ANOVA shows that all parameters were not statistically significant main effect of means at different time points for P ulse rate (p = Page of 0.994), Temperature (p = 0.9952), Systolic Blood Pressure (p = 0.9948) and Diastolic Blood Pressure (p = 0.9943).

Since the Hematopoietic parameters of Hemoglobin, WBC, Neutrophils, and Platelet Count were n ormally distributed and, T-Test was used to compare the means for Pre-Dose and Post-Dose. Whereas the rest of all other parameters i.e., RBC, Lymphocytes, Eosinophil, Monocytes, PCV were normall y distributed and, the Wilcoxon signed-rank-test used to compare the means for Pre-Dose and Post-Dose.

The results of using the T-Test revealed that there was no statistically significant difference between Means for Neutrophils (p = 0.4924), for WBC (p = 0.0543), for Platelet Count (p = 0.3584). Whereas the results of using the T-Test revealed that there was a statistically significant difference between Means for Hemoglobin (p = 0.0034).

The results of using the Wilcoxon signed-rank-test revealed that there was no statistically significant difference between Means for Lymphocytes (p = 0.9197), for Eosinophil (p = 0.2852), for Monocytes (p = 0.1514). Whereas the results of using the Wilcoxon signed-rank-test revealed that there was a statistically significant difference between means for RBC (p = 0.0008) and for PCV (p = 0.0005).

There was no adverse event observed, all organ system was normal in function so no need for any a dditional intensive investigation. IP Tolerances were considered in the study for the tolerability of investigati onal products. IP tolerance evaluation was performed on days 5, 6 and after 7 days of IP administration, After evaluation on days 5, 6 and after 7 days IP administration there was no side effect on any organ system.

### 3.2. Clinical trial phase 2

#### 3.2.a. Patients

Patients were randomized, 50 were assigned to receive ATRICOV 452 and 50 to receive placebo. Three patients in placebo and two patients in ATRICOV 452 group had dropped out from the study due to unknown reasons. Hence, the primary analysis population included 95 patients (48 allocated to ATRICOV 452 and 47 to placebo).

Patients in both groups were balanced in demographic and disease characteristics at baseline. The mean age of patients in the primary analysis population was 39 years (interquartile range [IQR], 22-59), 46 (52%) were women, and 44(48%) were men, they did not have any known comorbidities at baseline. The most common were, fever (94 patients, 98.94%) and cough (95 patients, 100%), followed by respiratory rate (baseline mean 13.16) and SPO2 (baseline mean 93.244).

#### 3.2.b. Primary Outcome

Time to resolution of symptoms in patients assigned to ATRICOV 452 vs placebo was not significantly different (median, 6 days vs 7 days). In the ATRICOV 452 and placebo groups, symptoms resolved in 100% of patients by day 15. The type of placebo that patients received did not affect the results.

Swab test results for all (100) subjects found positive at screening visit (visit 1) and after EOT (visit 9) test results performed for 95, and all 95 subjects were negative.

Subject wise data was observed from visit 1 to EOT and mean values of all COVID-19 specific inflammatory markers (NLR, CRP, Ferritin, D-dimer, IL-6, and PCT) levels were significantly improved compared to placebo group.

The mean percentage improvement in NLR levels on visits 6, 8, and 9 with reference to visit 1 were -28%, -24%, and -19% respectively in ATRICOV 452 treated group. Whereas in the placebo group the mean percentage improvement in NLR levels on visits 6, 8, and 9 with reference to visit 1 were 7%, 9%, and -5% respectively. This has illustrated the level of NLR was improved from a visit to visit while it was worsened in the placebo-treated group.

The mean percentage improvement in CRP levels on visits 6, 8, and 9 with reference to visit 1 were -76%, -75%, and -55% respectively in ATRICOV 452 treated group. Whereas in the placebo group the mean percentage improvement in CRP levels on visits 6, 8, and 9 with reference to visit 1 were -94%, -66%, and -36% respectively. This has illustrated the proportionate improvement in CRP levels from a visit to visit in both ATRICOV 452 and placebo treated groups. However, the mean CRP level during visit 1 was less in ATRICOV 452 treated group, it has further improved and the CRP level were normal during visit 8.

The mean percentage improvement in ferritin levels on visits 6, 8, and 9 with reference to visit 1 were -2%, -9%, and 0.5 % respectively in ATRICOV 452 treated group. Whereas in the placebo group the mean percentage improvement in CRP levels on visits 6, 8, and 9 with reference to visit 1 were -198%, -175%, and -17% respectively. This has illustrated the proportionate improvement in ferritin levels from a visit to visit in both ATRICOV 452 and placebo treated groups.

The mean percentage improvement in D-Dimer levels on visits 6, 8, and 9 with reference to visit 1 were -22%, -11%, and 2% respectively in ATRICOV 452 treated group. Whereas in the placebo group the mean percentage improvement in D-Dimer levels on visits 6, 8, and 9 with reference to visit 1 were -6%, -1%, and 4% respectively. This has illustrated the proportionate improvement in ferritin levels from a visit to visit in both ATRICOV 452 and placebo treated groups.

The mean percentage improvement in IL-6 levels on visits 6, 8, and 9 with reference to visit 1 were 27%, 30%, and 34% respectively in ATRICOV 452 treated group. Whereas in the placebo group the mean percentage improvement in IL-6 levels on visits 6, 8, and 9 with reference to visit 1 were 21%, 36%, and 40% respectively. This has illustrated the proportionate improvement in ferritin levels from a visit to visit in both ATRICOV 452 and placebo treated groups.

The mean percentage improvement in PCT levels on visits 6, 8, and 9 with reference to visit 1 were -34%, -24%, and 9% respectively in ATRICOV 452 treated group. Whereas in the placebo group the mean percentage improvement in PCT levels on visits 6, 8, and 9 with reference to visit 1 were -27%, -40%, and 0.2% respectively. This has illustrated the proportionate improvement in ferritin levels from a visit to visit in both ATRICOV 452 and placebo treated groups.

Considering exploratory nature and analysis of study outcomes, the safety and tolerability of ATRICOV 452 in improving the level of COVID-19 specific inflammatory markers in adult COVID-19 patients was assessed. No serious adverse events occurred during the study and ATRICOV 452 found well tolerated.

## 4. Conclusion

The investigational product, ATRICOV 452 capsule of 500 mg, has been found to be safe and well-tolerated for single dose study, without any safety concerns. The CTRI registered Phase 2 trial performed with two arms (Placebo & ATRICOV 452) has significantly reduced the level of COVID-19 specific inflammatory markers and no adverse events were reported.

## Data Availability

The data is archived, available at trial site and with the sponsor.

## 5. Acknowledgement

## References

1. WHO Coronavirus Disease (COVID-19) Dashboard accessed on 30/9/2020, 4:28pm CEST.

2. Sanders JM, Monogue ML, Jodlowski TZ, Cutrell JB. Pharmacologic Treatments for Coronavirus Disease 2019 (COVID-19): A Review. JAMA. 2020;323(18):1824–1836. doi:10.1001/jama.2020.6019.

3. Jason Phua, Li Weng, Lowell Ling, Moritoki Egi, Chae-Man Lim, Jigeeshu Vasishtha Divatia, et al., Intensive care management of coronavirus disease 2019 (COVID-19): challenges and recommendations. A Review. The Lancet Respiratory Disease, 2020, 8, 506–517.

4. Wang M, Cao R, Zhang L, Yang X, Liu J, Xu M, Shi Z, Hu Z, Zhong W, Xiao G. Remdesivir and chloroquine effectively inhibit the recently emerged novel coronavirus (2019-nCoV) in vitro. Cell Res. 2020 Mar; 30(3):269–271.

5. Li G, De Clercq E. Therapeutic options for the 2019 novel coronavirus (2019-nCoV). Nat Rev Drug Discov. 2020 Mar; 19(3):149–150.

6. Chen N, Zhou M, Dong X, Qu J, Gong F, Han Y, Qiu Y, Wang J, Liu Y, Wei Y, Xia J, Yu T, Zhang X, Zhang L. Epidemiological and clinical characteristics of 99 cases of 2019 novel coronavirus pneumonia in Wuhan, China: a descriptive study. Lancet. 2020 Feb 15; 395(10223):507–513.

7. Melika Lotfi,a b Michael R. Hamblin,c,d,e and Nima Rezaeif, COVID-19: Transmission, prevention, and potential therapeutic opportunities. Clin Chim Acta. 2020 Sep; 508: 254–266.

8. Latha Damle, Hrishikesh Damle, Shiban Ganju, Chandrashekar C, Bharath B R. In silico, In vitro Screening of Plant Extracts for Anti-SARS-CoV-2 Activity and Evaluation of Their Acute and Sub-Acute Toxicity. bioRxiv 2021.09.07.459230; doi: https://doi.org/10.1101/2021.09.07.459230

